# Prevalence of *Mycoplasma genitalium* infection and macrolide and fluoroquinolone resistance mutations among U.S. Air Force service members living with HIV, 2016-2020

**DOI:** 10.1101/2024.04.24.24306271

**Authors:** Shilpa Hakre, Eric Sanders-Buell, Rosemary O. Casimier, Anne Marie O’Sullivan, Sheila A. Peel, Sodsai Tovanabutra, Paul T. Scott, Jason F. Okulicz

## Abstract

**Background:** *Mycoplasma genitalium* (MG) infection is of public health concern due to antimicrobial resistance (AMR). Data are limited on repeat MG infection and AMR among United States Air Force (USAF) service members (SMs) living with human immunodeficiency virus (HIV).

**Methods:** USAF SMs seeking HIV care were screened for MG infection during the surveillance period (May 16, 2016-March 16, 2020). Baseline and repeat MG prevalence were estimated. An extended Cox proportional hazards regression model evaluated characteristics associated with repeat MG infection. MG-positive rectal samples were tested for macrolide or fluoroquinolone resistance.

**Results:** Among 299 male patients of a total 308 patients followed during the surveillance period, baseline prevalence of MG infection was 19.7% (59/299) and repeat MG was 35% (36/101) among patients who screened positive for MG at any time during the surveillance period. Characteristics independently associated with increased risk of repeat infection were reported prior sexually-transmitted infection (STI) history vs none (adjusted hazards ratio (aHR) 2.21, 95% confidence interval (CI) 1.13-4.35), presence of STI coinfection vs not indicated (aHR 5.13, 95% CI 2.78-9.49), new HIV diagnosis (<1 year vs 1 year or more, aHR 2.63, 95% CI 1.62-4.27). AMR in MG-positive rectal specimens was 88% (43/49) indicating macrolide resistance, 18% (10/56) with quinolone resistance, and 18% (10/56) with both macrolides and fluoroquinolone resistance.

**Conclusions:** Macrolide and fluoroquinolone resistance mutations were common. Testing for co-occurring MG infection and AMR mutations may be warranted in guiding treatment for STIs such as chlamydia or gonorrhea detected at HIV diagnosis.

## BACKGROUND

*Mycoplasma genitalium* (MG) is a sexually transmitted infection (STI) which has been linked to non-gonococcal urethritis (NGU), cervicitis, pelvic inflammatory disease, infertility and preterm birth.[1, 2] [3, 4] The prevalence of MG infection varies by country and by the population studied. In the United States (U.S.) and Britain, the prevalence among men and women in the general population has been low ranging from 1.0% to 1.7% whereas in less developed countries the summary prevalence was 3.9%.[5–8] In clinic-based studies, the prevalence ranged from 28.7% among men with urethritis in the U.S., 20.8% among asymptomatic women with bacterial vaginosis enrolled from ten U.S. cities, 16.6% among attendees of sexual health clinics in six U.S. cities, and 16.7% among HIV pre-exposure prophylaxis (PrEP) users in high income countries. [9–12] HIV infection can increase susceptibility to sexually transmitted infections (STIs), and potentially complicate treatment due to compromised immune function. Compared to uninfected men, men with HIV bear a reportedly six-fold higher overall burden of MG although this association was not observed for *Chlamydia trachomatis* and *Neisseria gonorrhoeae*.[13]

The emergence of macrolide-/fluorquinolone-resistant strains of MG complicated management using first-line therapy with macrolides (azithromycin) and second-line therapy with fluoroquinolones (moxifloxacin) in the U.S. leading to a change in treatment guidelines in 2021 [14, 15] Macrolide resistance leading to treatment failure has been associated with five single nucleotide mutations within the V region of the 23S rRNA gene at positions 2058 and 2059 (*Escherichia coli* numbering).[16] Resistance to fluoroquinolones has been linked to amino acid substitutions in the quinolone resistance-determining region (QRDR) of the gyrA and parC gene. A decline in moxifloxacin efficacy from 100% to 89% has been reported due to resistance associated with mutations found in gyrA/B and parC/E genes.[17]

Historically STI has posed a significant infectious disease threat to U.S. military personnel, potentially amplifying the problem of MG infection.[18] While some studies have investigated the prevalence of MG among U.S. military personnel with and without HIV, reports specifically focusing on antimicrobial resistance, repeat infection, and risk factors are absent.[19–22] Treatment failure due to antimicrobial-resistant MG and subsequent reinfection especially in an immunocompromised population could present a patient management and disease control problem deserving attention. This study sought to describe the prevalence of repeat MG infection among U.S. Air Force (USAF) service members with HIV, to identify associated factors, and to evaluate the prevalence of macrolide- and quinolone-associated resistance exhibited by MG isolates from follow-up of USAF service members who sought specialty care for HIV while in service.

## METHODS

### Patient population

USAF service members seeking routine HIV care were tested for MG and *Trichomonas vaginalis* (TV) in addition to a routine STI test panel including *Chlamydia trachomatis* and *Neisseria gonorrhoeae*. During the surveillance period, all USAF military personnel diagnosed with HIV were provided specialty care at an infectious disease clinic located in San Antonio, Texas. Details of collection of specimens and risk information have been reported previously.[21] Briefly, service members were administered a one-page questionnaire eliciting demographic, STI and sexual risk history. An extra pharyngeal and rectal swab, and urine specimen aliquot were collected for MG and TV send-out nucleic acid amplification testing (Aptima *M. genitalium* research-use-only analyte-specific reagents and Aptima *T. vaginalis*, Hologic) to the HIV Diagnostics and Reference Laboratory (HDRL, Walter Reed Institute of Research (WRAIR), Silver Spring, Maryland).

### Ethics

The Walter Reed Army Institute of Research Division of Human Subject Protection and the Defense Centers of Public Health-Aberdeen (formerly named Army Public Health Center) Public Health Review Board determined the project was a public health activity and not research.

### Antimicrobial resistance sequencing

Urine and extragenital specimens collected at the infectious disease clinic in San Antonio, Texas from May 16, 2016, through March 16, 2020 were placed in transport tubes (Hologic) prior to shipping to HDRL for diagnostic testing. The transport tube contained buffer which lysed target cells and released target ribosomal RNA thus preventing degradation during storage. Sample aliquots were frozen at -80 degrees Celsius until December 10, 2020, when all available aliquots were transferred to the Viral Sequencing Core laboratory (VSC, WRAIR) for sequencing. The main gene targets for macrolide – 23S - and fluoroquinolone - parC, and gyrA were selected for investigation, along with other targets around the L4 and L22 proteins of the ribosomal complex known to confer macrolide resistance in other molluscites such as *Mycoplasma pneumoniae*.[16, 23, 24]

DNA was extracted from available volume of all non-rectal swab samples ranging from 0.3 to 0.7 ml using DNA mini kit (QIAGEN, Germantown, Maryland). For rectal swab samples, DNA isolation was performed using the NucleoSpin DNA Stool, Mini kit for DNA (Macherey-Nagel Inc., Allentown, Pennsylvania) with an additional wash with ST4 buffer as recommended by the supplier. Nucleic acids were amplified using a nested PCR strategy with Titanium Taq (Takara Bio USA, Inc., San Jose, California) for each region. The amplification primers were as shown in Table 1. The primer sequences were either obtained from the literature for the regions 23S, L4 and L22, parC and Gyr, or designed using for *M. genitalium* strain 75956 (GenBank: CP145105) as a template for alternative amplification primer sites proximal to target regions.[16, 23, 25] Candidate primers were further subjected to a BLASTn search in order to maximize specificity to MG by excluding primers that had any similarity to other pathogens or human genes matched in the BLASTn search.

**Table 1.**
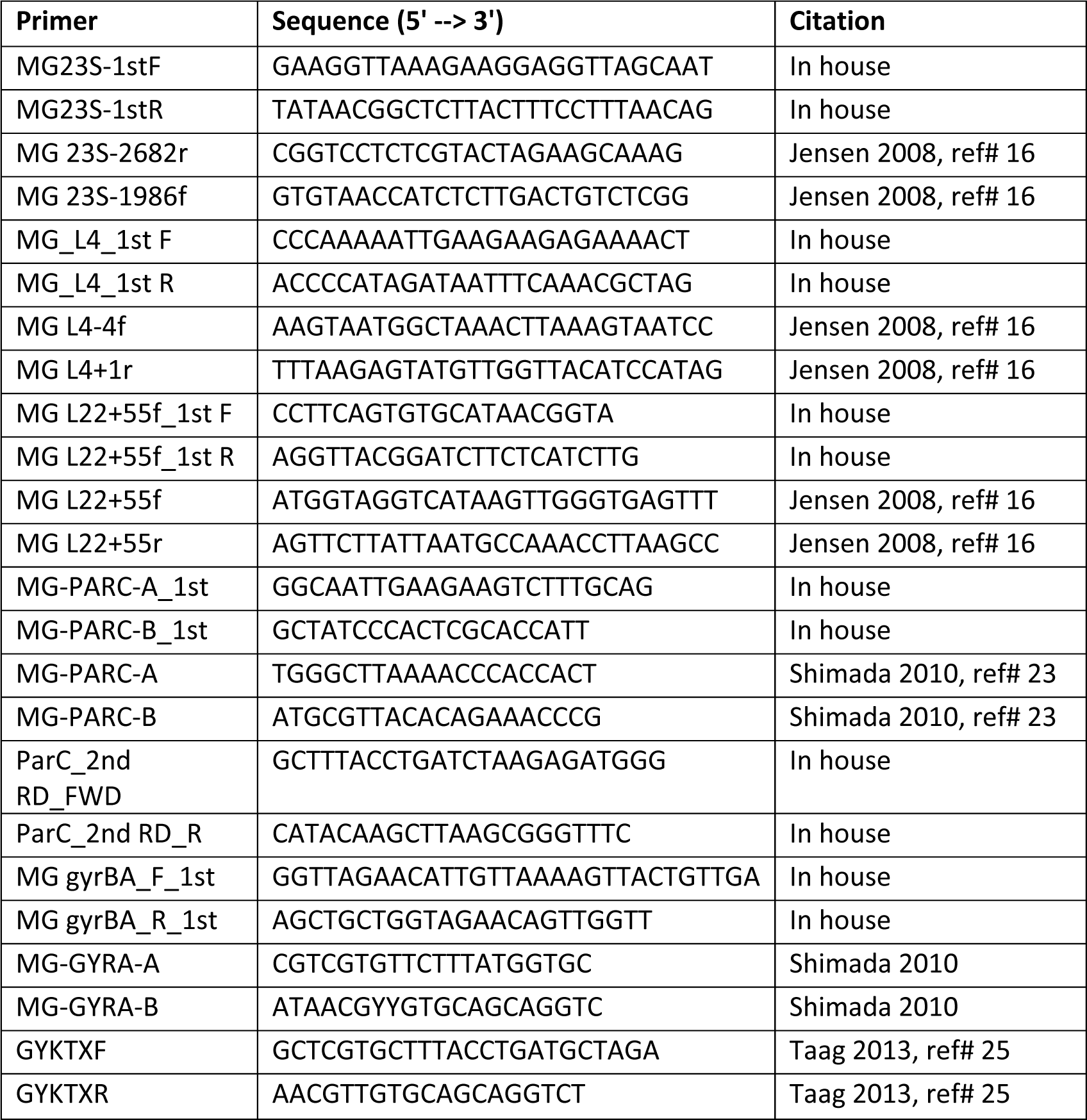
Primers used for sequencing of *Mycoplasma genitalium* in the study.

PCR cycling conditions for the various amplifications include standard denaturation of 95°C 30 sec and 68 °C 2 min extension for 30 cycles. The annealing conditions varied per each region as follow for 30 sec: 23S region 60°C 1st round 63°C and 2nd round, par C 55°C 1st round 58°C and 2nd round, L4 63°C 1st round 55°C and 2nd round. Gyrase and L22 annealing conditions were 60°C for both rounds of PCR. Samples that failed to amplify were then subjected to a post extraction genome amplification with Genomiphi (Cytiva ™, Marlborough, Massachusetts) prior to PCR. PCR cycling conditions for the various amplifications include standard denaturation of 95°C 30 sec and 68 °C 2min extension for 30 cycles. The annealing conditions varied per each region as follow for 30 sec: 23S region 60°C 1st round 63°C and 2nd round, Par C 55°C 1st round 58°C and 2nd round, L4 63°C 1st round 55°C and 2nd round. Gyrase and L22 annealing conditions were 60°C for both rounds of PCR.

Sanger sequencing was performed on the amplicon of the targeted regions directly on both strands using Big Dye terminator reaction kits and an ABI_3730xl genetic analyzer (ThermoFisher, Foster City, California). DNA sequences were assembled using Sequencher software version 3.1 (Genecodes Inc., Ann Arbor, Michigan). The obtained sequences were aligned to reference sequence for *M. genitalium* strain M2321 (GenBank: CP003770.1).

For macrolide resistance, nucleic acid changes were evaluated at position 2058 or 2059 for the 23S gene.[16] For fluoroquinolone resistance, mutations at amino acid positions AA 84, 87 or 97 Y of the parC gene (topoisomerase) were examined.[23]

### Statistical analysis

The proportion of patients with baseline prevalence, incident, repeat, and macrolide-/fluoroquinolone-resistant MG were described during follow up in the surveillance period. Baseline prevalence was assessed at the first visit whereas patients testing positive after having had a negative result were considered to have an incident infection. A repeat positive MG test result for any specimen type in patients diagnosed with MG at any time during the surveillance period was defined as a repeat MG infection. Descriptive statistical analyses were used to explore differences between demographic, behavioral, and laboratory characteristics at entry for patients with single versus repeat infection using the Chi-square, Fisher’s exact, or Kruskal-Wallis tests. All 95% confidence intervals (CI) were two-tailed and statistical significance was assessed at a p-value <0.05. The Andersen-Gill extension of the Cox proportional hazards regression model was fit to the data to evaluate baseline and time-varying patient characteristics associated with repeat MG.[26] All patient characteristics were self-reported except for STI coinfection status which was extracted from electronic medical records. All data management and analyses were performed using SAS version 9.2 (SAS Institute, Cary, North Carolina).

## RESULTS

A total of 2,417 specimens were collected and tested from 308 patients who received infectious disease specialty care for HIV over the course of four years from May 16, 2016 through March 16, 2020. Analysis in this report was restricted to male patients (n=299, 97%), since only nine patients were female. Among females who were excluded, one patient’s MG-positive rectal specimen was identified with macrolide resistance (A2059G, L22 bp81G>R). *T. vaginalis* was excluded from analysis since only eight infections were identified among men in the surveillance period.

Male patients were followed a median 1.8 years (interquartile range (IQR) 0.5-2.8) and tested a median 9 (IQR 6-12) times accounting for each specimen type. Most patients (198, 66%) had no diagnosis of MG infection in the surveillance period, whereas almost a quarter (65, 22%) were diagnosed with infection only once (Figure 1). Patients detected with a single infection were followed a median 2.0 years (IQR 0.6-2.7) which did not differ significantly from those detected with infection more than once (median 2.1, IQR 1.1-3.0, p>0.05). Baseline prevalence of MG was 19.7% (59/299, 95% CI 15.5-24.5) and was highest for rectal (13.9%, 41/294) specimens, followed by urine (6.7%, 18/268), and pharyngeal (1.7%, 5/296) specimens. Among 240 patients who tested negative for MG at their first surveillance visit, 47 were diagnosed with MG infection during follow up for an incidence rate of 12.1 per 100 person-years (95% CI 8.9-16.1). Among an overall 101 (33.8%) patients diagnosed with MG infection at any time during the surveillance period, 36 (35%) were detected with MG repeatedly up to 5 times for a repeat diagnosis rate of 47 events per 100 person-years (95% CI 33.7-64.8). Repeat infections were identified a median 11.9 months (IQR 6.9-12.6) apart.

**Figure 1.**
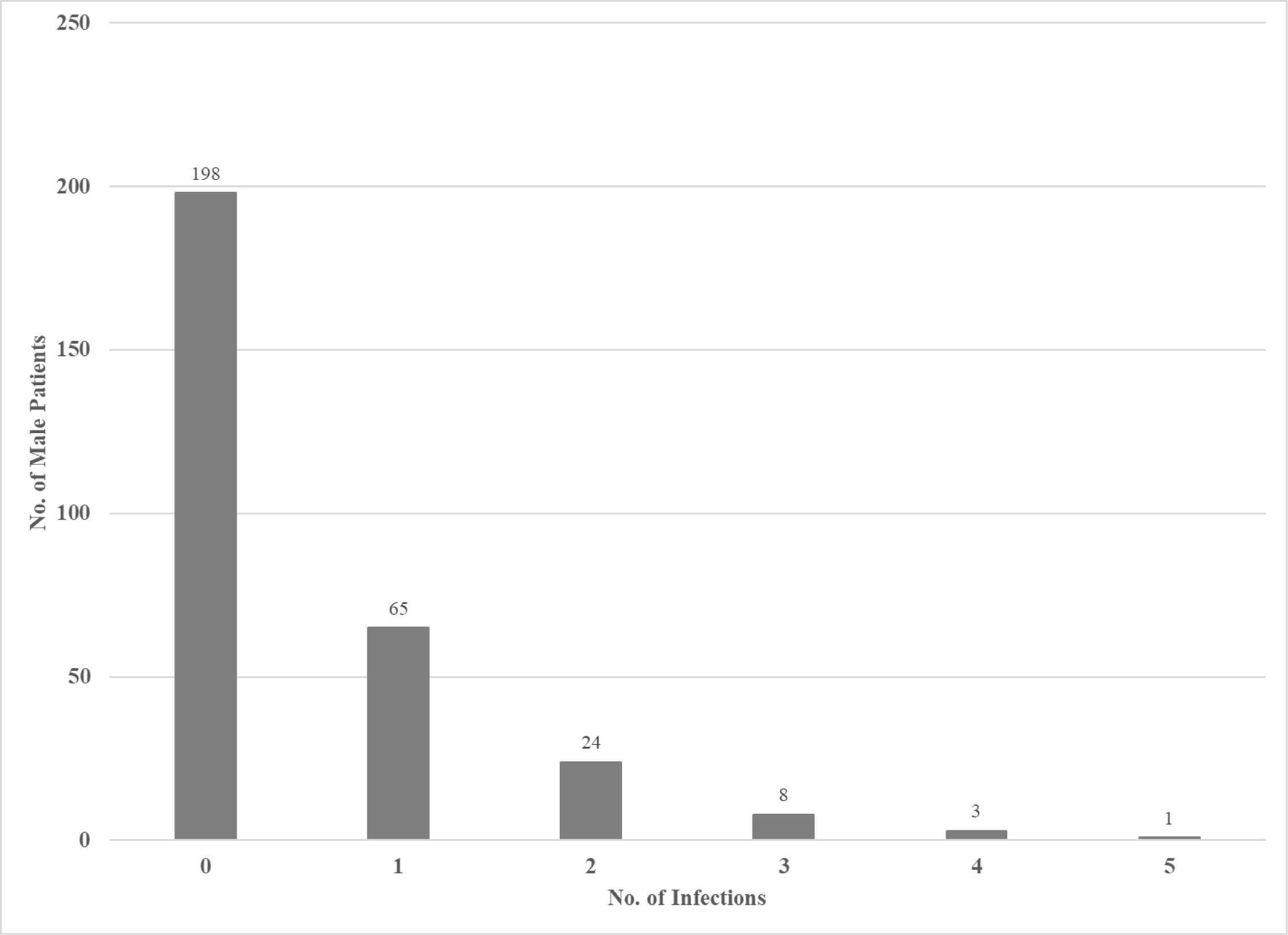
Male U.S. Air Force patients with HIV diagnosed with *Mycoplasma genitalium*, May 16, 2016 through March 16, 2020.

Most male patients were 19-39 (90%) years of age, enlisted (84%), and single (57%) (Table 2). A higher proportion (25%) of patients with repeat infections reported prior STI symptoms at entry compared to those with a single infection (12%) (Table 2). In univariate analysis, three characteristics were associated significantly (p<0.05) with increased risk of repeat infection (Table 2). These were self-report of prior STI (vs none, HR 4.00, 95% CI 1.87-8.56), having an STI coinfection (vs not indicated, HR 5.58, 95% CI 2.41-12.9), and a recent (<1 year) diagnosis of HIV (vs 1 year or more, HR 3.56, 95% CI 1.72-7.37). Although risk decreased after adjustment in multivariate analysis, the three characteristics remained independently associated with increased risk of repeat infection (reported prior STI history vs none, adjusted HR (aHR) 2.21, 95% CI 1.13-4.35; presence of STI coinfection vs not indicated, aHR 5.13, 95% CI 2.78-9.49; <1 year since HIV diagnosis vs 1 year or more, aHR 2.63, 95% CI 1.62-4.27) (Table 3).

**Table 2.**
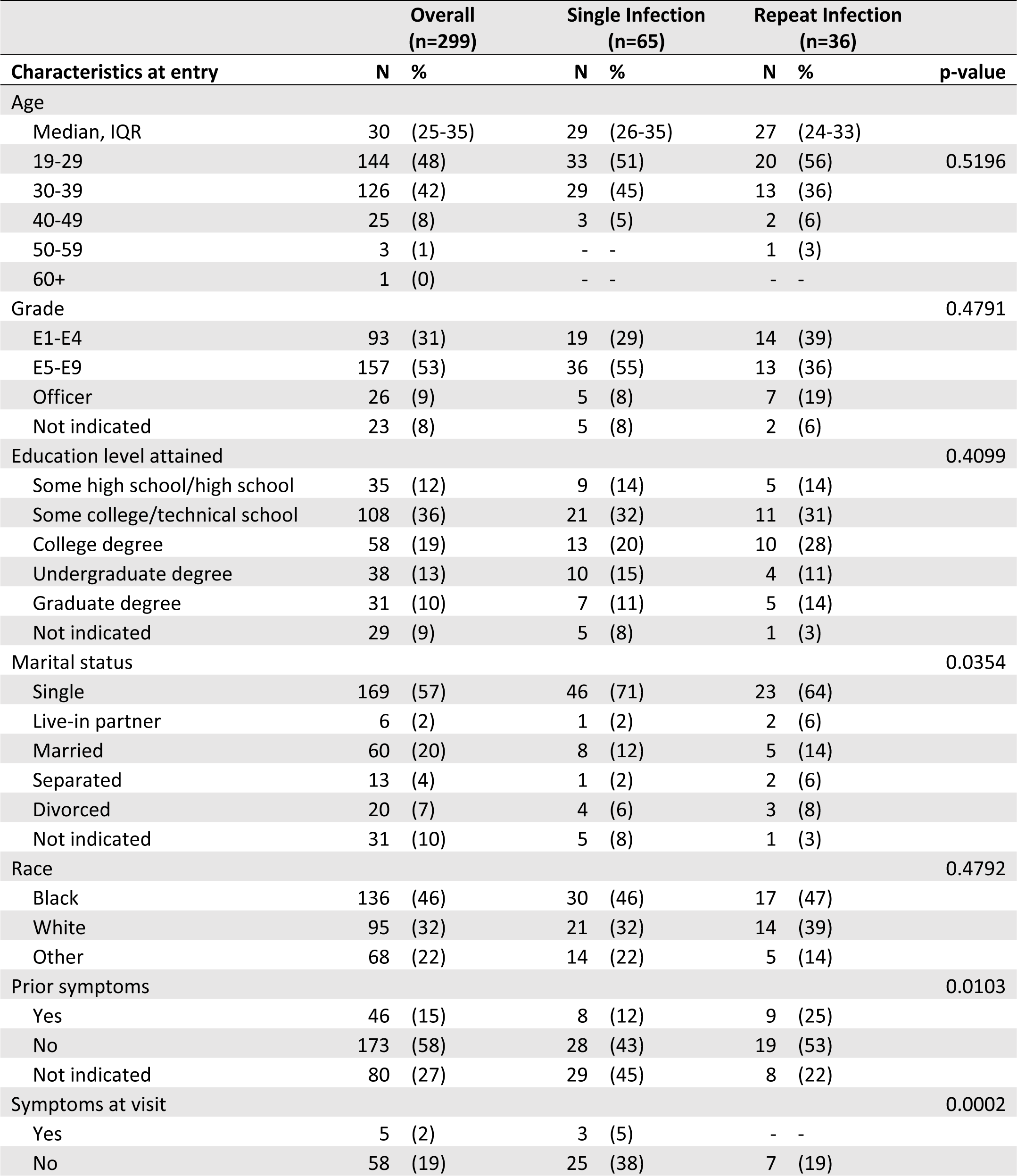
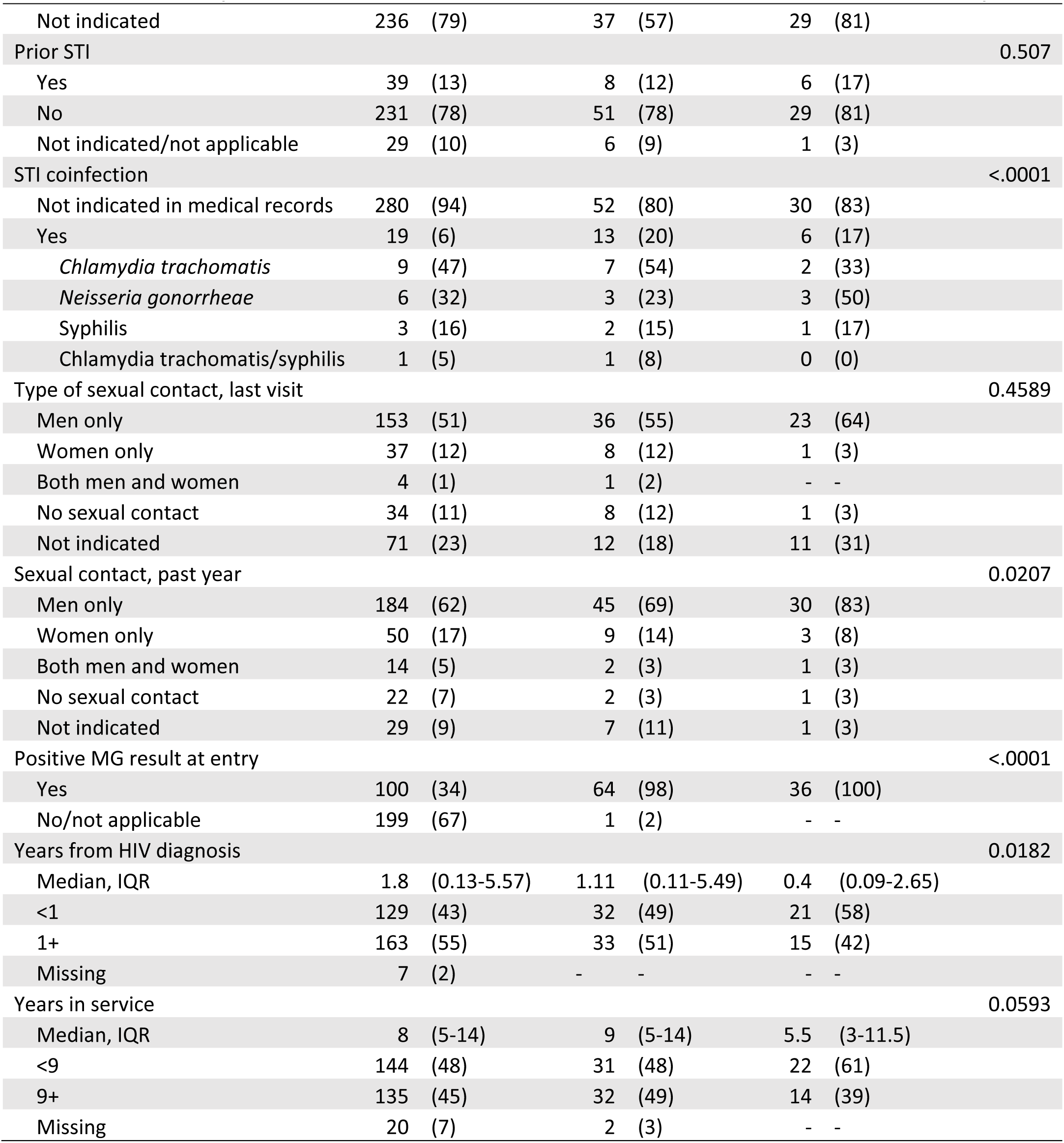
Sociodemographic, clinical, and sexual behavior characteristics of 299 male Air Force service members with HIV, overall, and by *Mycoplasma genitalium* positivity, 2016-2020.

**Table 3.** Unadjusted and adjusted hazard ratios (HR) for male Air Force service members living with HIV with specimens repeatedly positive for *Mycoplasma genitalium*, 2016-2020.

| Characteristic | Comparison | Unadjusted HR | 95% CI | p-value | Adjusted HR | 95% CI | p-value |
| --- | --- | --- | --- | --- | --- | --- | --- |
| Age at entry | 19-29 vs 30+ | 1.36 | (0.65-2.86) | 0.41 |  |  |  |
| Age at sample collection | 19-29 vs 30+ | 1.24 | (0.60-2.57) | 0.56 |  |  |  |
| Race | White vs black/other | 1.98 | (1.00-3.91) | 0.05 |  |  |  |
| Paygrade | E5-E9 vs E1-E4 | 0.52 | (0.23-1.17) | 0.11 |  |  |  |
|  | Officer vs E1-E4 | 1.23 | (0.36-4.19) | 0.75 |  |  |  |
| Highest education attained | Some college/technical school vs high school or less | 0.35 | (0.11-1.07) | 0.07 |  |  |  |
|  | College degree vs high school or less | 0.48 | (0.15-1.56) | 0.22 |  |  |  |
|  | Undergraduate degree vs high school or less | 0.37 | (0.08-1.67) | 0.20 |  |  |  |
|  | Graduate degree vs high school or less | 0.71 | (0.20-2.52) | 0.59 |  |  |  |
| Marital status | Single vs ever married | 1.16 | (0.51-2.65) | 0.73 |  |  |  |
| STI symptoms since last visit | Yes vs no | 1.86 | (0.75-4.64) | 0.18 |  |  |  |
| Prior STI history | Yes vs no | 4.00 | (1.87-8.56) | 0.0004 | 2.33 | (1.26-4.31) | 0.007 |
| STI coinfection | Yes vs not indicated | 5.58 | (2.41-12.9) | <.0001 | 5.13 | (2.78-9.49) | <.0001 |
| Type of sexual partner since the last visit | Men vs women/both men and women/no sex | 1.46 | (0.61-3.53) | 0.40 |  |  |  |
| No. of male sexual partners since last visit | 1 or more vs none | 1.87 | (0.24-14.87) | 0.55 |  |  |  |
| No. of new male sexual partners since last visit | 1 or more vs none | 1.47 | (0.67-3.22) | 0.34 |  |  |  |

| Characteristic | Comparison | Unadjusted HR | 95% CI | p-value | Adjusted |  |  |
| --- | --- | --- | --- | --- | --- | --- | --- |
|  |  |  |  |  | HR | 95% CI | p-value |
| No. of female sexual partners since last visit | 1 or more vs none | 0.16 | (0.02-1.00) | 0.05 |  |  |  |
| Type of sexual partner in the past year | Men vs women/both men and women/no sex | 3.68 | (0.75-18.03) | 0.11 |  |  |  |
| No. of new male sexual partners in the past year | 1 or more vs none | 1.18 | (0.55-2.52) | 0.68 |  |  |  |
| No. of female sexual partners in the past year | 1 or more vs none | 0.22 | (0.04-1.33) | 0.10 |  |  |  |
| Years since HIV diagnosis at study entry | <1 vs 1+ | 3.56 | (1.72-7.37) | 0.0006 | 2.33 | (1.45-3.73) | 0.0004 |
| Years in service at study entry | <9 vs 9+ | 1.69 | (0.80-3.56) | 0.17 |  |  |  |
| Years in service at sample collection | <9 vs 9+ | 1.59 | (0.78-3.25) | 0.20 |  |  |  |
| Macrolide sensitivity | Resistant vs sensitive | 1.12 | (0.30-4.17) | 0.87 |  |  |  |
| Quinolone sensitivity | Resistant vs sensitive | 1.25 | (0.42-3.70) | 0.69 |  |  |  |

Note: Confidence intervals (CI)

Among 56 rectal specimens from 43 patients sequenced for evidence of antibiotic resistance to MG, 88% (43/49) had markers indicating macrolide resistance, 18% (10/56) indicating quinolone resistance, and 18% (10/56) had resistance markers to both macrolides and quinolones. The most common macrolide-associated mutations which were detected were A2058G (n=21) and A2059G (n=18) (Table 4). Other uncommon mutations were A2058R, A2058T, A2059C (n=2), and A2059R (n=1).

**Table 4.**
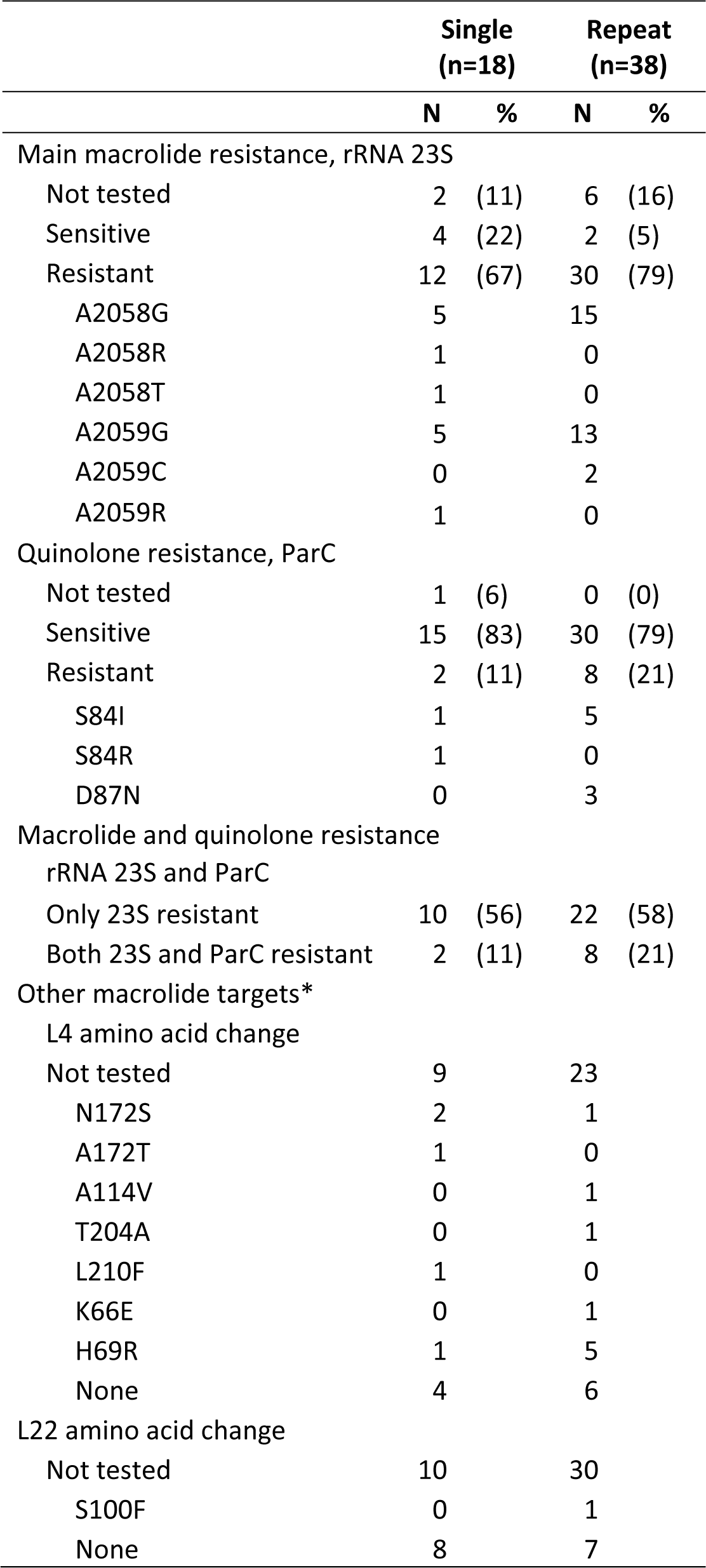
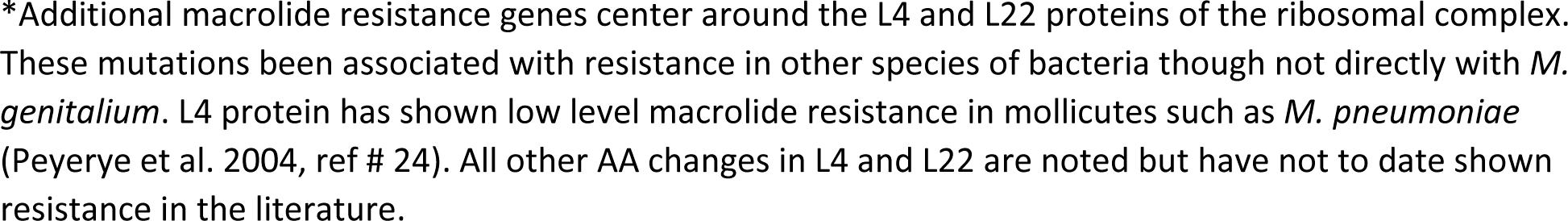
Proportion of 56 rectal samples positive for *Mycoplasma genitalium* and sequenced for macrolide and/or fluoroquinolone resistance mutations among Air Force service members with human immunodeficiency virus diagnosed with single or repeat MG infection, 2016-2020.

|  | Single<br>(n=18) |  | Repeat<br>(n=38) |  |
| --- | --- | --- | --- | --- |
|  | N | % | N | % |
| Main macrolide resistance, rRNA 23S |  |  |  |  |
| Not tested | 2 | (11) | 6 | (16) |
| Sensitive | 4 | (22) | 2 | (5) |
| Resistant | 12 | (67) | 30 | (79) |
| A2058G | 5 |  | 15 |  |
| A2058R | 1 |  | 0 |  |
| A2058T | 1 |  | 0 |  |
| A2059G | 5 |  | 13 |  |
| A2059C | 0 |  | 2 |  |
| A2059R | 1 |  | 0 |  |
| Quinolone resistance, ParC |  |  |  |  |
| Not tested | 1 | (6) | 0 | (0) |
| Sensitive | 15 | (83) | 30 | (79) |
| Resistant | 2 | (11) | 8 | (21) |
| S84I | 1 |  | 5 |  |
| S84R | 1 |  | 0 |  |
| D87N | 0 |  | 3 |  |
| Macrolide and quinolone resistance<br>rRNA 23S and ParC |  |  |  |  |
| Only 23S resistant | 10 | (56) | 22 | (58) |
| Both 23S and ParC resistant | 2 | (11) | 8 | (21) |
| Other macrolide targets* |  |  |  |  |
| L4 amino acid change |  |  |  |  |
| Not tested | 9 |  | 23 |  |
| N172S | 2 |  | 1 |  |
| A172T | 1 |  | 0 |  |
| A114V | 0 |  | 1 |  |
| T204A | 0 |  | 1 |  |
| L210F | 1 |  | 0 |  |
| K66E | 0 |  | 1 |  |
| H69R | 1 |  | 5 |  |
| None | 4 |  | 6 |  |
| L22 amino acid change |  |  |  |  |
| Not tested | 10 |  | 30 |  |
| S100F | 0 |  | 1 |  |
| None | 8 |  | 7 |  |

\*Additional macrolide resistance genes center around the L4 and L22 proteins of the ribosomal complex. These mutations been associated with resistance in other species of bacteria though not directly with *M. genitalium*. L4 protein has shown low level macrolide resistance in mollicutes such as *M. pneumoniae* (Peyerye et al. 2004, ref # 24). All other AA changes in L4 and L22 are noted but have not to date shown resistance in the literature.

Quinolone-associated mutations detected for ParC were AA84 S to I (n=6) and S to R (n=1), and AA87 D to N (n=3). Among 43 patients whose samples were sequenced, more than half (58%) had repeat infections. Twenty one of 43 patients had more than one sample sequenced. A higher proportion of samples with resistance were from repeat infections, especially for macrolides (repeat 78% vs single infection 63%) (Table 4). No samples were found to be subsequently resistant to macrolides or fluoroquinolones after initial sensitivity. Patient characteristics did not differ significantly by age, grade, marital status, and education for those whose samples were tested for resistance compared to other MG-positive patients who were not tested (p>0.05).

## DISCUSSION

The prevalence of MG infection was 18.6% among the first 102 Air Force service members with HIV included in a prior analysis of this enhanced surveillance project. In this follow-up analysis which included an additional 197 male service members with HIV, a similar baseline prevalence (19.7%) of MG was identified. The incidence rate was 12.1 cases per 100 person-years among service members who tested negative at baseline. Macrolide or quinolone resistance was commonly identified (88% and 18%, respectively) among almost half of the patients with MG-positive specimens that were sequenced.

Although studies of MG infection among men living with HIV are limited, estimates in this analysis are comparable to those reported from other developed countries. Prevalence and incidence of MG were 20.3% and 19.5 cases per 100 person-years, respectively, among 148 men who have sex with men (MSM) with primary HIV screened in a prospective study in Zurich, Switzerland, and 21% among 48 MSM with HIV who sought care at STI or HIV clinics for proctitis in Melbourne, Australia; incidence was not assessed in the latter study. [27, 28]

Recent diagnosis of HIV was associated with repeat MG infection in this analysis suggesting a link to a weakened immune system. In one review, MG prevalence was found to be highest among acquired immune deficiency syndrome (AIDS) patients (43.7%) with a markedly lower burden among asymptomatic persons living with HIV (27.3%) and uninfected individuals (11.3%).[29] In other studies, MG infection has been bi-directionally associated with HIV, found to increase shedding of HIV DNA, and thought to facilitate HIV transmission in women by compromising the epithelial lining of the urogenital tract and increase susceptibility of peripheral blood mononuclear cells to HIV.[29–32] The role of MG infection in men is unclear although it is postulated that inflammation of the rectal mucosa due to MG infection may increase presence of HIV-susceptible cells in men without HIV and facilitate HIV acquisition.[27]

In this analysis, self-reported prior STI history and STI coinfection were associated with repeat MG infection. Coinfection with other STIs or infection with a single STI has been reported to increase the risk of a subsequent STI or to increase the risk of HIV.[30, 33, 34] Furthermore, these, and other factors such as recent macrolide use, and male-to-male sexual contact have been reportedly associated with macrolide- and/or fluoroquinolone-associated resistance mutations.[34–37] Moreover, MSM with recurrent STIs had a higher prevalence of MG infections and antibiotic resistance-associated mutations. It’s been suggested that the higher prevalence of mutations may be due to increased chance of antimicrobial selection pressure from treatment of co-occurring STIs such as chlamydia or gonorrhea in high STI risk populations such as MSM.[34] Single dose azithromycin was part of the recommended treatment for chlamydia or gonorrhea infection before a change in guidelines in 2021 to treating non-pregnant individuals with doxycycline for 7 days.[15, 38]

Resistance to macrolides or fluoroquinolones in this analysis were 88% and 18%, respectively, among 43% of patients with MG whose samples were sequenced. These estimates are comparable to the prevalence of 68.4-90.5% reported among high risk populations such as clinic attendees seeking care for NGU, cervicitis, proctitis, and MSM seeking STI testing.[39, 40] [34, 41] Globally macrolide and fluoroquinolone resistance in MG increased at least five-fold in a decade. The overall prevalence of macrolide resistance across 21 countries increased from 2010 (10%) to 2016-2017 (51.4%) whereas the prevalence of resistance associated with fluoroquinolone (7.7%) did not vary markedly over time. [42] Comparatively, the prevalence of macrolide resistance in the Americas increased even more from 0% to 67.3% in the same period. Similarly, the prevalence of fluoroquinolone resistance reported in the World Health Organization (WHO)-Americas region was higher at 10·1% for the period 2013–2017.

A limitation of this analysis was samples from less than half of patients with detected MG were sequenced due to resource limitations or sample unavailability. However, the preliminary finding of a higher proportion of repeat infection samples having resistance in combination with the association of STI history and coinfection with repeat MG suggest treatment of prior or concurrent bacterial STI infections may play a role in development of resistance in MG strains. Alternatively, resistant MG strains may have been sexually transmitted from an infected sexual partner.

In this first report evaluating resistant strains of MG among U.S. military service members with HIV, prevalence of macrolide- and/or fluoroquinolone resistance was commonly found in rectal specimens, especially for repeat positive samples. New diagnosis of HIV, STI history and/or co-occurrence were associated with the detection of repeat MG. Further research is needed to fully understand the complex interplay of factors contributing to repeat MG diagnosis in men living with HIV and whether testing for co-occurring MG infection and antibiotic resistance mutations may be warranted in guiding treatment for STIs such as chlamydia or gonorrhea found at HIV diagnosis.

## Data Availability

To protect the privacy of individuals currently serving in the military, data are not publicly available. For inquiries, please contact the WRAIR Public Affairs Office at

## NOTES

## Acknowledgements

We thank Elizabeth J. Bianchi and Erifile Zografos for data capture; Catherine Stewart for diagnostic testing and procurement; Meera Bose, Morgan DiGiorgio, Rachel Loney, Kelly Parisi, and Meghan Waters for sequencing; and Joanna Freeman and Christopher Schoening for laboratory data exports.

## Disclaimer

Material has been reviewed by the Walter Reed Army Institute of Research. There is no objection to its presentation and/or publication. The opinions or assertions contained herein are the private views of the author, and are not to be construed as official, or as reflecting true views of the Department of the Army or the Department of Defense, Ministry of Health (Jordan), or the Henry M. Jackson Foundation for the Advancement of Military Medicine, Inc. (HJF). The investigators have adhered to the policies for protection of human subjects as prescribed in AR 70–25.

